# Relationship between midwifery density and midwifery regulatory environments in International Confederation of Midwives member countries

**DOI:** 10.1101/2024.07.02.24309863

**Authors:** Emma Virginia Clark, Marianna LaNoue, Kate Clouse, Alexandra Zuber, Jeremy Neal

## Abstract

**Background:** Midwives are a cost-effective solution for reducing preventable maternal death, but barriers to their recruitment, training, deployment, and retention exist. Improved midwifery regulation is proposed to address these challenges through activities related to education and qualification, licensure, registration/re-licensure, and scope and conduct of practice. However, no quantitative analysis of the associations between midwifery regulation and workforce density in low- and middle-income countries has been conducted, limiting actionable insights for policymakers. The objective of this study was to evaluate the relationship between midwifery regulatory environments and midwifery workforce densities (midwives per 10,000 population) in International Confederation of Midwives member countries.

**Methods:** The study used secondary data from the 2019 Global Midwives’ Associations Map Survey to perform a cross-sectional analysis of 103 International Confederation of Midwives member countries. To operationalize the strength of a country’s midwifery regulatory environment, we used a previously developed Midwifery Regulatory Environment Index. We conducted multivariable linear regression analysis to evaluate the relationship between this index and country-level midwifery density, available from the National Health Workforce Accounts.

**Results:** Midwifery regulatory environments are significantly associated with midwifery workforce densities across countries. For every 1-point increase in the Midwifery Regulatory Environment Index (indicating activities in at least one additional regulatory domain), midwifery density increased by 0.66 midwives per 10,000 population (p = 0.011). This effect decreased to 0.41 midwives per 10,000 population yet remained significant even after controlling for country level income, per capita health spending, income inequality, and human development index (p = 0.04).

**Conclusion:** Stronger regulatory environments are associated with higher densities of midwives, even when controlling for potential confounding factors such as country-level income and health spending. This relationship suggests that policymakers should invest in midwifery regulation to improve maternal health outcomes.

## Introduction

The current global sexual, reproductive, maternal, newborn, and adolescent health (SRMNAH) workforce is insufficient, meeting only 75% of the global need for essential services [1]. This workforce provides care across the reproductive lifespan, including preconception (e.g., offering modern contraceptive methods), early pregnancy (e.g., post-abortion care, ectopic pregnancy management), antenatal (e.g., screening and managing hypertensive disorders, diabetes, and malaria), intrapartum (e.g., parenteral administration of anticonvulsants, uterotonics, and antibiotics, manual removal of placenta), and postpartum (e.g., promoting breastfeeding) periods [2]. In low-income countries, only up to 41% of the need for these essential services is met [1].

Midwives are increasingly recognized as a cost-effective means for delivering SRMNAH services that improve outcomes and reduce preventable maternal and newborn mortality [3–5], yet an estimated 900,000 more midwives are needed to meet global service delivery needs [1]. The availability of a country’s health workforce to provide essential SRMNAH services in any given geographical area can be estimated using national workforce density reports, with low- and middle-income countries (LMICs) having the greatest workforce shortfall [6]. Numerous regulatory barriers at the country level can contribute to this shortfall, hindering the recruitment, training, deployment, and retention of midwives. These health professional regulation barriers include unclear and non-standardized role definitions, limited scopes of practice misaligned with international standards, varying educational and clinical standards for entry to practice, and weak licensure and registration authorities and systems [7,8].

Health professional regulation establishes mechanisms and standards that recognize and delineate the roles of health professionals, providing flexibility to adapt to complex and evolving health system contexts [9]. According to the World Health Organization (WHO), regulation is a priority component of advancing the health workforce, and policymakers should consider it a strategic tool for addressing workforce imbalances and other challenges to ensure adequate training and availability of health professionals [10]. Specifically, midwifery regulation defines criteria and processes outlined in legislation to identify what a midwife is and to describe their scope of practice [11]. The aim of regulation is to promote regulatory mechanisms that protect the public (women and families) by ensuring that safe and competent midwives provide high standards of midwifery care to every woman and baby through such actions as producing autonomous, high-quality practice [11].

The presence of distinct regulatory structures and processes for midwifery is crucial to enable autonomous midwifery practice and maintain midwifery standards, both of which are essential for providing high-quality care to mothers and infants [11]. Midwifery regulation can be categorized into the following regulatory domains, which are derived from the International Confederation of Midwives (ICM) Global Standards for Midwives [11] and adapted through additional research [12–14].

- **Overarching Regulatory Policy and Legislation:** Enacting legislation that recognizes midwifery as a profession distinct from nursing and establishing a dedicated regulatory body with specifically for midwives or with separate and distinct policies and processes for midwives.
- **Education and Qualification**: Establishing standards for midwifery education, curricula, and, occasionally, accreditation of educational institutions to develop necessary competence for practice.
- **Licensure:** Developing and enforcing criteria, standards, and processes for initial entry into practice.
- **Registration/Re-Licensure**: Establishing continuing medical education or other requirements for demonstrating continued competence and maintaining a current database of midwives, including their registration/licensure status, to support workforce planning, and consumer transparency.
- **Scope and Conduct of Practice:** Clearly defining the scope of practice of a midwife consistent with ICM’s definition of a midwife and establishing mechanisms for handling complaints and disciplinary action.

A strong midwifery regulatory environment is one in which midwifery is recognized as a distinct profession, and a specific regulatory body, led by midwives, exists. Alternatively, a distinct track for midwifery regulation within a nursing board or council may suffice. This empowered body, established through regulatory policies, has statutory authority and effectively implements and oversees activities across the domains noted above. Conversely, a weak regulatory environment is characterized by the absence, inadequacy, or non-functionality of regulatory activities and mechanisms.

Midwifery density is the number of midwives per 10,000 population in a specified geographic area. Density data are critical for assessing workforce availability, recruitment, deployment, and retention. The WHO and global professional associations have emphasized the significance of regulation in advancing the midwifery workforce, resulting in a growing body of research aiming to formally establish linkages between regulation and midwifery workforce densities. Predominantly conducted in high-income countries, these studies have explored how regulations constraining midwifery practice can lead to a reduction in the midwifery workforce [15–17].

Research in the United States has underscored that strong regulatory environments are related to higher midwifery workforce densities [18,19]. However, limited evidence exists on this relationship in LMICs, where research mostly focuses on documenting midwifery workforce challenges and suggesting that improved regulation might alleviate these challenges, without exploration of the underlying mechanisms. For example, a scoping review in Kenya [20] and a descriptive review in Nigeria [21] effectively highlight how poor job satisfaction and low retention rates contribute to midwifery shortages, with the suggestion that weak regulatory environments may be a driving factor. However, similar to other LMIC-focused research, these studies lacked a quantitative analysis of associations between regulation and workforce, limiting actionable insights for policymakers.

This study aims to evaluate the relationship between midwifery regulatory environments and midwifery workforce densities in ICM member countries, addressing a critical gap in the literature. We hypothesize that countries with stronger midwifery regulatory environments will have higher densities of midwives.

## Methods

This study was conducted using a cross-sectional design and secondary data analysis of multiple publicly available databases. Approval for this study was obtained from the Vanderbilt University Medical Center Institutional Review Board.

### Data and Variables

The independent variable for this study was the Midwifery Regulatory Environment (MRE) Index, a composite score our team developed to operationalize the strength of a country’s midwifery regulatory environment. Data used in developing this index were drawn from the ICM Global Midwives’ Association Map Survey. For this survey, ICM collected data from country midwifery associations with support from the United Nations Population Fund (UNFPA) Country Office and relevant government organizations between October and December 2019 [22]. The survey included four subsections on midwifery: association, education, leadership, and regulation. To ensure data quality, ICM used a multi-step process including an initial review with country follow-up for clarifications and completeness, internal logic, and validity; review of the revised and validated data by country midwifery associations; and a letter of validation from an official for each survey section or confirmation that data were publicly available from reputable sources [22].

To develop the MRE Index, we began by identifying relevant data from the Map Survey for inclusion in the composite score. The regulatory sub-section contained 38 questions assessing a country’s regulatory environment [23]. We eliminated sixteen questions for free-text answers and/or not directly relating to at least one regulatory domain. The remaining 22 questions were consolidated and organized into 12 items, each associating with one of the five essential domains noted above [overarching regulatory policy and legislation (4 items); education and qualification (0 items); licensure (4 items); registration/re-licensure (2 items); and scope and conduct of practice (2 items)]. Because the education and qualification domain was not reflected in the regulatory sub-section questions, one question from the education subsection of the Map Survey about the national curriculum for midwifery education was included as a final item for MRE Index development purposes. This resulted in a total of 13 items for inclusion in calculation of the final composite MRE Index.

Five different potential scoring methodologies for aggregating the 13 items into a single MRE Index were developed, and the characterization that best fit the data and had the highest predictive ability for each of three outcomes (low birthweight prevalence, stillbirth rate, MMR) was selected. While all characterizations performed well, the “Aggregated Domain Scoring” characterization proved to be the best. In this approach, the 13 items are grouped into five categories, each representing a regulatory domain: overarching regulatory policy and legislation, education and qualification, licensure, registration/re-licensure, or scope and conduct of practice. One point is assigned to each domain with at least one “yes” response to the item(s) within that domain, allowing for a total score between zero (no regulatory activities in any domain) and five (regulatory activities in all five domains). Detailed methodology regarding development of the MRE Index is published in a forthcoming manuscript.

The dependent variable for this study was the density of midwives at the country level, defined as the number of midwives per 10,000 population. These data were obtained from WHO’s National Health Workforce Accounts (NHWA), which facilitates data reporting on key workforce indicators to improve the availability, quality, and use of health workforce data with the goal of achieving universal health coverage [24]. WHO’s 194 member states submit health workforce data annually to the NHWA. These data sources include health workforce registries or databases, aggregated data from health facilities such as routine administrative records and District Health Information System census or surveys, professional council, chamber, and/or association registers, and labor force surveys [25]. The NHWA uses population estimates provided by the United Nations (UN) Statistics Division, drawn from civil registration and vital statistics systems, population censuses or registers, population registers, and household surveys. If other methodologies are used, WHO recalculates densities using the UN Statistics Division population estimates described above to harmonize densities [25].

Covariate data were obtained from publicly available databases including the United Nations Development Program’s (UNDP) Human Development Index (HDI) [26] and WHO’s Global Health Expenditure database [27], and the World Bank’s Gini index [28] and income group databases [29]. Four control variables were selected to reduce potential bias related to overall allocation and distribution of resources within countries. These included two continuous variables: per capita health spending in United States dollars and the Gini index, which measures the extent to which the distribution of income or consumption among individuals or households within an economy deviates from a perfectly equal distribution [28]. We also used two categorical variables. A country’s income group is assigned annually by the World Bank based on the gross national income (GNI) per capita of the previous year, expressed in United States dollars. In 2022, countries were classified as low income (GNI per capita less than $1,085), lower-middle income (GNI per capita $1,086-4,255), upper-middle income (GNI per capita $4,256-13,205), and high income (GNI per capita greater than $13,205) [29]. The HDI, the final control variable, is a composite measure of average achievement in three key dimensions of human life: a long and healthy life measured by life expectancy at birth; education, measured by mean years of schooling and expected years of schooling; and a decent standard of living, measured by gross national income per capita in Purchasing Power Parity (PPP) international dollars. It is reported on a scale from 0 to 1.00, with ≥ 0.800 classified as “very high” human development, 0.700-0.799 as high, 0.550-0.699 as medium, and < 0.550 as low [26].

### Data Screening and Analysis

One hundred fifteen countries submitted complete responses to the regulatory subsection of the ICM Global Midwives’ Association Map Survey, which was used for calculating the MRE Index. Three countries were missing density information, and nine others were missing data for one or more covariates, leaving a total of 103 countries for analyses. We transformed two highly-skewed variables, midwifery workforce density and per capita health spending, by selecting the positive square root transformation which had the best effect on skewness and kurtosis. We used multivariable linear regression to test for associations between the MRE Index and midwifery density, adjusting for the covariates. Statistical significance was set *a priori* at 0.05. SPSS Statistics 27 was used to conduct the analysis.

## Results

MRE Indices across countries ranged from 2 to 5, with an average score of 4.36 (SD = 0.88). Midwifery workforce density ranged from 0.11 midwives per 10,000 population in Burundi to 20.46 midwives per 10,000 population in Ireland. The median density was 2.41 midwives per 10,000 population (interquartile range [IQR] 1.43, 5.10). Table 1 presents descriptive summaries of MRE Index, midwifery workforce density, and control variables used in the regression analysis (i.e., per capita health spending, Gini index, income group, and HDI).

**Table 1.** Descriptive statistical summaries of country characteristics (N = 103)

|  | Mean | SD |
| --- | --- | --- |
| **MRE Index** | 4.36 | 0.88 |
| **Gini Index (1-100)** | 37.39 | 7.48 |
|  | Median | IQR |
| **Health Spending (USD)** | 201.06 | 58.5, 1278.18 |
| **Density of Midwives (per 10,000 population)** | 2.41 | 1.43, 5.10 |
|  | % |  |
| **Human Development Index** |  |  |
| Low | 22.0 |  |
| Medium | 28.0 |  |
| High | 17.0 |  |
| Very High | 33.0 |  |
| **Income Level** |  |  |
| Low | 25.0 |  |
| Low-Middle | 29.0 |  |
| Upper-Middle | 17.0 |  |
| High | 29.0 |  |

Correlations among the explanatory variables indicate that all control variables were significantly associated with each other, with health spending, Gini index, and income group demonstrating very strong associations (Table 2). However, MRE Index was not significantly associated with any of the covariates.

**Table 2:** Correlations among MRE Index and control variables (N= 103)

|  | MRE Index | Health Spending<sup>a</sup> | Income Group | Human Development Index |
| --- | --- | --- | --- | --- |
| Health Spending<sup>a</sup> | -0.097 | - |  |  |
| Income Group | 1.19 | 155.037*** | - |  |
| Human Development Index | 1.42 | 101.55*** | 181.055*** | - |
| Gini Index | 0.003 | 0.304* | 9.35*** | 8.10*** |

Chi square for categorical variables, Person r for continuous variables, one-way ANOVA for mixed categorical (>2 categories) & continuous variables.
\* = $p < 0.05$ , \*\* = $p < 0.01$ , \*\*\* $p < 0.001$ .
<sup>a</sup> transformed variable

Results of univariate and multivariate regression analyses are shown in Table 3. The multiple regression of all explanatory variables with midwifery workforce density was significant (Multiple *R* = 0.405, p = 0.010), explaining approximately 12% of the variance. While all explanatory variables showed significant univariate correlations with midwifery workforce density, only health spending and the MRE Index remained significant after adjusting for health spending, HDI, Gini index, and income level.

**Table 3:** Summary of univariate and multivariate associations with midwifery density (N = 103)

| Characteristic | *r* | *p*-value | *beta*<sup>c</sup> | SE | 95% Wald Confidence Interval |  | *p*-value |
| --- | --- | --- | --- | --- | --- | --- | --- |
|  |  |  |  |  | Lower | Upper |  |
| MRE Index | -0.24 | 0.011 | 0.41 | .013 | -0.37 | 1.19 | 0.04 |
| Health Spending (USD) | 0.31 | <0.001 | 0.00 | 0.00 | 0.00 | 0.00 | 0.03 |
| Income Group<sup>a</sup> | 5.00 | 0.003 |  |  |  |  |  |
| 2-1 |  |  | -1.51 | 1.39 | -4.24 | 1.23 | 0.28 |
| 3-1 |  |  | -0.93 | 1.89 | -4.63 | 2.77 | 0.28 |
| 4-1 |  |  | 0.39 | 2.15 | -3.82 | 4.61 | 0.39 |
| Human Development Index<sup>b</sup> | 6.24 | <0.001 |  |  |  |  |  |
| 2-1 |  |  | 3.89 | 1.43 | 1.09 | 6.68 | 0.05 |
| 3-1 |  |  | 2.16 | 1.92 | -1.61 | 5.92 | 0.59 |
| 4-1 |  |  | 1.46 | 1.97 | -2.39 | 5.31 | 0.97 |
| Gini Index | 0.29 | 0.002 | -0.02 | 0.05 | -0.12 | 0.09 | 0.12 |

Multiple R = 0.405, $p = 0.010$ ; $R^2 = 0.164$ (Adjusted $R^2 = 0.120$ )
<sup>a</sup>Income Group: 1 = Low; 2 = Lower Middle; 3 = Upper Middle; 4 = High; all income group values are relative to low-income group.
<sup>b</sup>Human Development Group: 1 = Low; 2 = Middle; 3 = High; 4 = Very High.
<sup>c</sup>Beta obtained from model with untransformed variables for interpretation purposes.

Since transformed variables were used for midwifery workforce density and per capita health spending, *beta* values were calculated from non-transformed variables to aid interpretation. These values demonstrate that for every 1-point increase in the MRE Index (indicating activities in at least one additional regulatory domain), midwifery density increased by 0.66 midwives per 10,000 population (p = 0.011). This effect decreased to 0.41 midwives per 10,000 population when accounting for health spending, income group, inequality, and HDI, yet the MRE Index remained significantly associated with midwifery workforce density (p = 0.04).

## Discussion

This study aimed to determine if there is a relationship between midwifery regulatory environments and density of midwives in ICM member countries. The findings support our hypothesis that there is a positive association between the midwifery regulatory environments and midwife density; stronger regulatory environments are associated with higher densities.

This relationship remains significant even when controlling for potential confounding health system and country factors. The sample included a relatively balanced representation of low-, lower-middle, upper-middle, and high-income countries, as well as countries with varying levels of human development, although there were slightly more high-income and high human development countries.

Research such as the WHO’s “Midwives’ Voices, Midwives’ Realities” consultation highlights numerous issues that affect density. These issues include inadequate education, uncertainty about midwifery’s role in the health system, restrictions on practice that are inconsistent with the legal scope of practice, and overwork leading to burnout and attrition [30]. Each regulatory domain -- overarching regulatory policy and legislation, education and qualification, licensing, registration/re-licensure, and scope and conduct of practice – contains aspects that can influence midwifery density, with significant cross-over and reinforcement between domains. For example, in the overarching regulatory policy and legislation domain, Mattison et al. found when health system decision-makers did not recognize midwifery as an autonomous profession, such as through distinct regulatory bodies, midwifery was excluded from maternal health-related political agendas [31]. This omission led to downstream impacts such as lack of clarity about midwives’ specific roles in the health system and inappropriate deployment and remuneration. These factors significantly affect the desirability of midwifery as a profession and influence how many people choose to enter and remain in the profession.

Regulation in the education and qualification domain helps standardize education requirements for midwives, ensuring they meet at least minimum education standards and are prepared for their defined scope of practice. Aligning these standards with international norms further strengthens credibility, making the profession more attractive and drawing in more and stronger candidates. Administrative aspects of regulation, such as licensure and registration, can improve density by providing data on the number of midwives working in the country, distinguishing between active, inactive, and retired midwives, and identifying their locations.

This information is crucial for forecasting supply and analyzing it alongside demand data to understand labor market dynamics and gaps. Such insights support appropriate investments in midwifery training institutions and efforts to expand midwifery scope of practice, ensuring an adequate number of midwives are available to provide needed services. Lastly, activities within the scope and conduct of practice domain are fundamental aspects of regulatory environments [12] and major drivers of midwifery workforce density. Well-defined scopes of practice that promote strong interprofessional collaboration have been shown to attract and retain healthcare workers by clarifying task division, enhancing support, and managing overall workload [32,33]. A qualitative systematic review including studies from 23 LMICs found that workplace relationships between healthcare professionals that are built on trust contribute to social interactions and cooperation among healthcare workers, impacting their intrinsic motivation, retention, performance, and quality of care [34]. There is also evidence suggesting that midwives who practice autonomously – defined as the authority to make decisions and act freely based on their knowledge base– experience increased occupational satisfaction, improved professional identity, and reduced burnout [4,35,36]. Qualitative research on policymakers’ and regulators’ perspectives from both strong and weak regulatory environments could identify additional domains or domain activities not considered in this research. Research tracking health systems that are in the process of enhancing regulatory mechanisms for midwives, such as Lao People’s Democratic Republic [37], could also be evaluated using approaches that account for the complexity of health systems, such as difference-in-differences. It is also critical to ascertain if the relationship between midwifery workforce density and regulation holds at a sub-national level, as remote and rural areas often have much lower densities than urban areas and national averages, leading to poorer maternal, newborn, and child health outcomes [38]. While the NHWA has the capacity to report sub-national densities, only two countries in the study sample reported this information. As sub-national density data becomes more available, or in countries where it is currently available, this analysis could influence overall and specific domain investment in midwifery regulation.

### Limitations

This study had several limitations. As a secondary data analysis, it is limited by the quality of the datasets used. The data quality and validation processes for the main data sources are discussed, with quality variation being a generally accepted limitation of secondary data sources. Our regression analyses only included countries that are ICM members and participated in the ICM mapping survey, potentially introducing bias toward countries with more midwives and more sophisticated regulatory systems. However, many countries which are not ICM members do not have a midwifery cadre, mitigating some of this bias. Additional research is also needed to demonstrate a causal relationship between regulatory environments and midwifery workforce density to maximize policymaker interest in regulatory investments. Our findings only state an association, not causation, and these associations may be influenced by other factors in the health system or country governance, such as other investments in maternal and newborn health (e.g., abolition of user fees for MNH services).

## Conclusion

Stronger midwifery regulatory environments, characterized by regulatory policies and legislation, education and qualification standards, licensure, registration/re-licensure processes, and well-defined scopes and conduct of practice, are associated with higher densities of midwives globally. This likely results from the positive impact of regulation on the profession’s ability to attract, develop, and retain midwives due to enhanced professional identity, autonomy, and credibility. These findings underscore the importance of investing in midwifery regulation as a critical component for ensuring sufficient midwifery workforce and, consequently, improved maternal, newborn, and child health outcomes. However, additional research is needed to better understand how specific regulatory factors influence the pipeline of midwifery students, as well as the recruitment and retention of midwives, to establish causality and identify the role individual domains play in driving density.

## Data Availability

All relevant data are within the manuscript and its Supporting Information files.

https://apps.who.int/nhwaportal/

https://www.globalmidwiveshub.org/

